# Map Liberator: An open-source tool for recovering spatial epidemiological data from static situation reports

**DOI:** 10.64898/2026.01.26.26344575

**Authors:** David Simons

## Abstract

**Background:** Much of the world’s historical and current epidemiological data remains locked in static formats, such as PDF situation reports or image-based surveillance bulletins. Recovering this data for spatial analysis typically requires proprietary software (e.g., ArcGIS) or laborious manual entry, which is prone to transcription errors.

**Methods:** I developed Map Liberator, an open-source application built in R and Shiny. It provides a split-screen digitisation interface that allows users to overlay static reference maps alongside interactive administrative boundaries. The tool uses a stateful rendering engine to manage complex geometries (up to Admin Level 3) and facilitates the extraction of binary, numeric, or qualitative data directly into a structured, machine-readable format.

**Results:** I demonstrate the tool’s utility by digitizing Lassa Fever surveillance data from Nigeria (2018-2025). The tool successfully aligned static situation reports with official Local Government Area (LGA) boundaries, allowing for the rapid recovery of binary presence/absence data that was previously inaccessible for computational modelling.

**Conclusions:** Map Liberator provides a low-barrier, cost-effective solution for field epidemiologists and researchers to liberate spatial data from static reports, supporting the principles of FAIR (Findable, Accessible, Interoperable, Reusable) data in global health.

## Introduction

Effective epidemiological modelling and the design of targeted public health interventions increasingly require data at fine geographic scales. Whether coordinating vaccination campaigns, modelling vector-borne disease suitability, or identifying clusters of transmission, national or provincial-level aggregates are often insufficient for precision public health (Dowell, Blazes, and Desmond-Hellmann 2016; Tatem 2017; Bedford et al. 2019). However, obtaining data at the necessary sub-national resolution (e.g., district or municipality level) remains a significant challenge. Due to valid concerns regarding individual privacy, data ownership, or provisional reporting standards, granular surveillance data is frequently aggregated in public-facing reports (Gorina et al. 2020). Consequently, the most detailed spatial information is often presented solely as static choropleth or dot-density maps within PDF situation reports, rather than as accessible, machine-readable tables (Van Panhuis et al. 2014).

While there is a growing momentum towards FAIR (Findable, Accessible, Interoperable, Reusable) data standards in public health, exemplified by the European Centre for Disease Prevention and Control (ECDC) West Nile Virus dashboards, which provide both tabular and mapped data, adoption remains uneven (Wilkinson et al. 2016; García-Closas et al. 2023; European Centre for Disease Prevention and Control 2026). A substantial volume of historical and current epidemiological intelligence remains locked within static document formats (e.g., (Nigeria Centre for Disease Control and Prevention n.d.)). This reliance on static mapping is likely to persist during the transition period toward fully digital surveillance ecosystems, particularly in resource-constrained settings or during rapid-response scenarios where PDF bulletins remain the standard for dissemination.

Recovering data from these static maps for secondary analysis presents a methodological bottleneck. Current solutions typically fall into three categories, each with distinct limitations. First, proprietary Geographic Information Systems (GIS) software often includes georeferencing tools, but these operate within closed ecosystems that restrict access and reproducibility. Second, open-source alternatives (such as QGIS) offer robust functionality but possess steep learning curves, requiring specialist GIS knowledge that field epidemiologists or modellers may not possess (Kirby, Delmelle, and Eberth 2017). Third, manual transcription, visually cross-referencing a map and typing names into a spreadsheet, is laborious, unscalable, and introduces substantial need for post-processing to correct spelling errors or match non-standard place names to official geocodes (Goldberg, Wilson, and Knoblock 2007).

To address these limitations, I developed Map Liberator, an open-source tool designed to bridge the gap between static reports and spatial analysis (Simons [2025] 2026). Originally conceived to reconstruct historical cases of Lassa fever in Nigeria from weekly situation reports (2018–2025), the software has been generalised to be location - and disease - agnostic. Unlike traditional georeferencing tools that treat maps as raster images to be warped, Map Liberator focuses on the attribution of official administrative boundaries (Database of Global Administrative Areas 2022). By providing a streamlined, split-screen interface, the tool allows users to rapidly assign binary, numeric, or qualitative metadata to standard geometries.

This tool aims to improve data equity by lowering the technical barrier to accessing spatial data. Enabling practitioners and researchers to liberate data from static formats without requiring proprietary software or advanced GIS skills. While developed for infectious disease surveillance, its utility extends to any domain where data is visualised but not tabulated, facilitating the recovery of critical datasets for modelling and decision-making.

## Design and Implementation

### Software Architecture

Map Liberator is developed using the R statistical programming language (v4.2.3) and the Shiny web application framework (v1.11.1) (R Core Team 2023; Chang et al. 2025). I adopted a modular architecture to ensure maintainability and scalability, separating the application logic into distinct components: the map engine (mod_map_engine), the control interface (mod_controls), the reference viewer (mod_sidecar), and the data ledger (mod_workbench). This modularity facilitates future community contributions, allowing developers to enhance specific components (e.g., adding new data extraction modes) without disrupting the core application state. This modularity also supports long-term utility; it decouples the data source (GADM) from the application logic, ensuring that administrative boundaries can be updated via the setup.R script as new versions of the GADM database are released, without requiring a rewrite of the core engine.

### Data Preprocessing Pipeline

To ensure optimal performance within a browser-based environment, the application relies on a dedicated preprocessing pipeline (setup.R). Executed prior to deployment, this script programmatically retrieves administrative boundaries from the GADM database using the geodata package (Hijmans et al. 2024). It performs two critical operations: first, it harmonises the varying attribute schemas of different countries into a unified data structure, ensuring the application remains location-agnostic. Second using the terra package (v1.8-6) and rmapshaper (v0.5), high-resolution boundaries are simplified to reduce vertex density (Hijmans 2025; Teucher and Russell 2023). This substantially reduces file size while preserving topological integrity, ensuring that national-scale geometries (e.g., Nigeria’s 774 districts) render rapidly without inducing latency in the user interface. The processed data is stored as serialised R objects (.rds), balancing load speeds with memory efficiency.

### Stateful Spatial Engine

A significant challenge in web-based mapping tools is the computational cost of rendering complex geometries. To address this, I implemented a stateful rendering engine. This logic separates the initialisation of the map structure (tiles, panes, and z-indexes) from the data layer (polygons). When a user adjusts the map layout, such as expanding the map for detailed inspection, the application utilises a non-destructive JavaScript resizing method rather than re-rendering the elements. Furthermore, the engine uses leafletProxy to update vector styles (e.g., highlighting a selected district) in real-time without reloading the underlying geospatial data (Cheng et al. 2024).

### Split-Screen Digitisation Interface

A core utility of Map Liberator is its comparison interface. The application features a dynamic split-view that renders a static reference image alongside the interactive map. This sidecar module allows users to upload static image files (e.g., PNG/JPG). It includes a CSS-based transformation tools allowing the user to rotate and zoom the source map to align its orientation with the interactive canvas.

To facilitate the identification of boundaries within dense maps, the map engine implements an adaptive visual hierarchy. Administrative boundaries are rendered with varying stroke weights, thickest for national borders (Admin 0) and thinnest for Admin 3 boundaries, preventing visual clutter when working at fine spatial scales.

To expedite digitisation, the interface supports flexible data entry modes. Users can batch select multiple administrative units that share a common attribute — such as binary disease presence or identical case counts — and commit them to the ledger simultaneously. Conversely, for heterogeneous data (e.g., distinct numeric values per district), regions can be added individually. The application’s data state is decoupled from the active map view. This design allows users to clear the spatial viewer and load a new country or administrative level while retaining previously extracted records in the ledger. This capability enables the construction of transnational datasets (e.g., an outbreak spanning West Africa) within a single session.

Crucially, extracted data is not stored as a raster overlay but is immediately joined to official administrative names and unique identifiers (GIDs). This ensures that the output is not just a visual copy, but a semantic dataset ready for joining with other epidemiological variables or demographic datasets.

### Results (Validation Case Study)

#### Case Study: Lassa Fever in Nigeria (2018–2025)

To demonstrate the practical utility of Map Liberator, I performed a comprehensive extraction of Lassa fever case notifications from weekly Situation Reports published by the Nigeria Centre for Disease Control (NCDC). This extraction was necessitated by a specific research requirement: validating a multi-species occupancy modelling framework designed to predict Lassa mammarenavirus (LASV) risk across West Africa at a 5x5 km resolution. Standard public datasets provided only country-level aggregates, which were insufficient for validating such a high-resolution spatial model.

Accessing granular, district-level (Local Government Area, or LGA) data was therefore essential to accurately assess model performance. Data Extraction Process

I retrieved seven years of epidemiological surveillance data (2018–2025) from the NCDC archives. The relevant map displaying LGA attack rates (typically Figure 3 in the NCDC reports) was cropped from each PDF report. The extraction followed a rapid batch-processing workflow:

1. Nigeria Admin 2 boundaries (774 LGAs) were loaded into the map engine once at the start of the session.
2. A cropped map image was uploaded to the reference panel.
3. For each image, the project metadata (e.g., Year, Week) was updated within the control panel (Project Metadata and Data Attributes) (Figure 1.).
4. LGAs indicated as reporting cases were selected on the interactive map and coded as a binary variable (cases_detected = 1).
5. These data were added to the Data Ledger, and a new image was uploaded. The boundaries were cleared to reset the selections and steps 3 and 4 were repeated for each subsequent year, with the metadata updated to reference the data source and reference year. This allowed for the continuous compilation of a longitudinal dataset within a single session.

**Figure 1:**
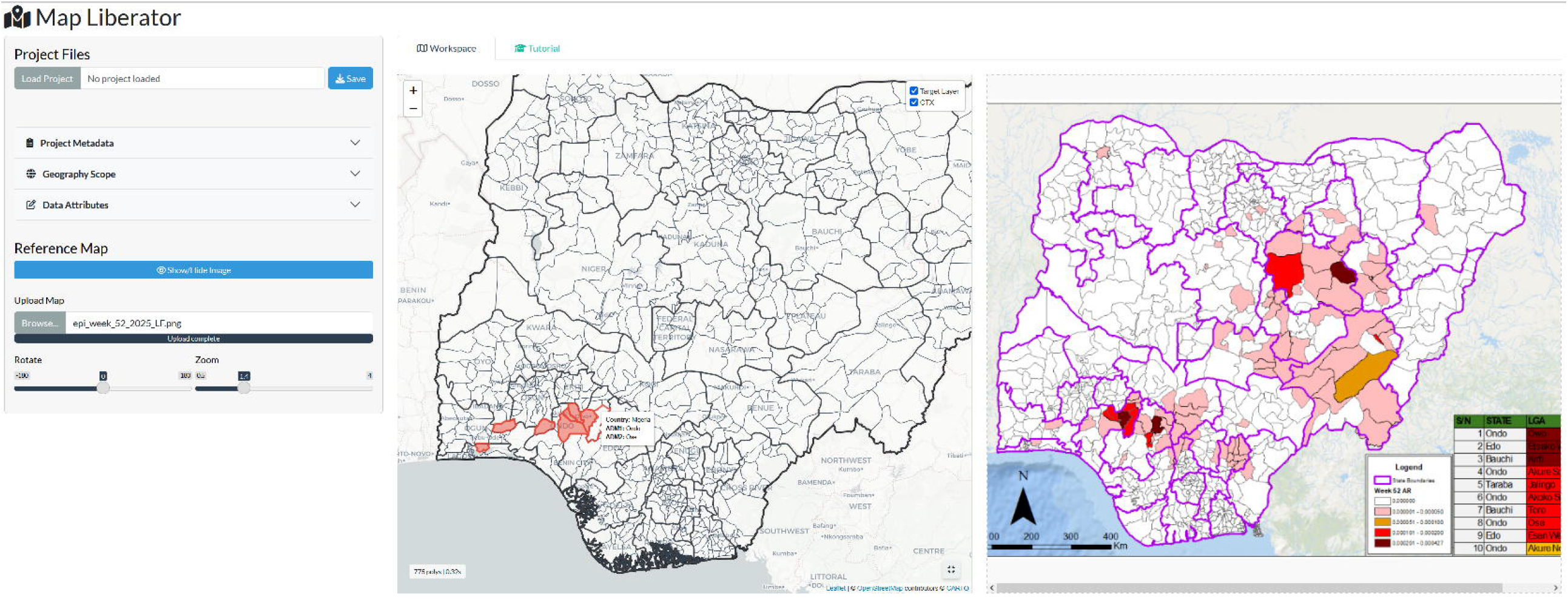
The split screen interface showing the reference NCDC map and interactive selection in a web browser. Red filled polygons represent selected LGAs which will be coded cases_detected = 1 in the data ledger

**Figure 2:**
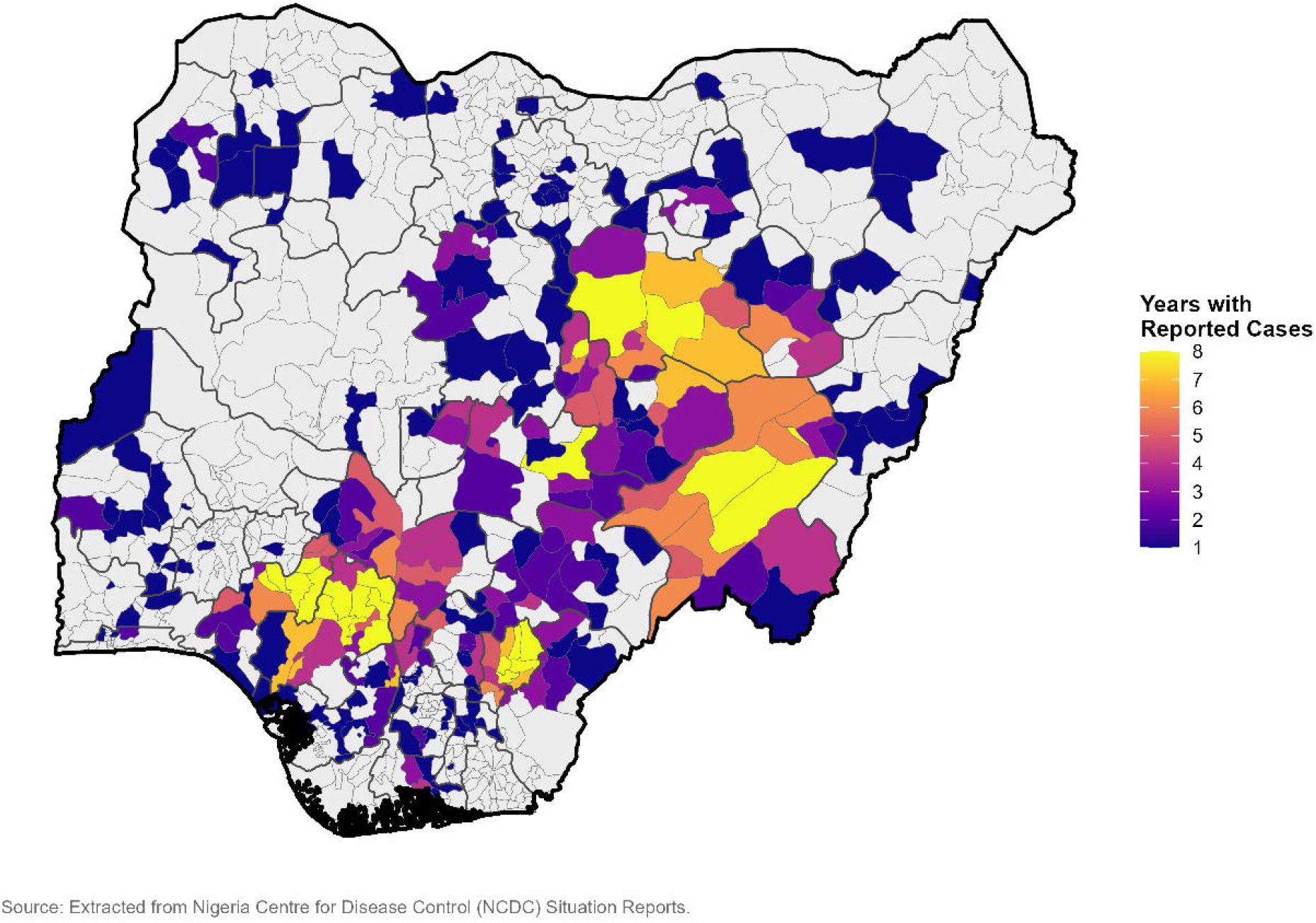
Spatial distribution of Lassa fever persistence in Nigeria (2018–2025). Shading represents the frequency of detection, defined as the number of years in which at least one confirmed case was reported in a Local Government Area (LGA). Data was extracted from weekly (week 52 of each year) NCDC Situation Reports using Map Liberator. Dark purple regions indicate sporadic reporting, while yellow regions indicate hyper-endemic areas reporting cases in all 8 years.

#### Outcome and Efficiency

The tool successfully generated a structured spatial dataset covering the seven-year period. The digitisation process averaged less than five minutes per map image. This represents a quantifiable improvement over the primary alternative: manual transcription. Traditional methods require a user to visually cross-reference a static map against an external gazetteer, identify the region name, and manually type data into a spreadsheet, a process that is not only slower but prone to typographic errors (e.g., misspelling ‘Esan West’). By utilising the interactive ‘click-to-assign’ interface, Map Liberator eliminates transcription errors entirely and ensures 100% linkage to valid administrative identifiers.

## Discussion

The FAIR compliant transition of epidemiological data is underway; however, critical surveillance intelligence remains accessible only as static images. Integrating these data remains a barrier to precision public health. Map Liberator addresses this challenge by providing a standardised, open-source workflow for recovering spatial data. To ensure the tool remains functional as R and Shiny evolve, Map Liberator relies on stable, widely-maintained dependencies (sf, terra, leaflet) rather than experimental packages. The open-source nature of the project (hosted on GitHub) allows the user community to submit issues and pull requests, ensuring continuous maintenance. Furthermore, the reliance on the programmatic retrieval of GADM data ensures that the tool can grow to support new administrative divisions without manual curation of shapefiles by the developer.

As demonstrated in the Nigeria case study, the tool significantly accelerates the digitisation process compared to manual transcription. By removing the need to visually cross-reference maps with external gazetteers, the risk of transcription errors (e.g., misspelling LGA names) is minimised. Furthermore, the ability to generate longitudinal datasets in a single session allows researchers to rapidly construct time-series baselines for modelling, as evidenced by the successful validation of the Lassa fever occupancy model (Simons 2025).

The tool’s accuracy is contingent on two factors: the legibility/resolution of the source map and the quality of the underlying administrative boundaries. Map Liberator currently relies on GADM data. While GADM is a global standard, it may not always reflect the most recent local administrative changes (e.g., newly created districts) or historical administrative districts. Users working in countries with frequent boundary reorganisations may encounter discrepancies between the static report and the interactive map. I have identified this as a potential future update to the process where users can upload their own shapefiles which can be incorporated into the application.

## Data Availability

All data used in the case study are available on the Nigeria Centre for Disease Control and Prevention website. The extracted data have been made available as supplementary information. Software code is available on GitHub and an archived version is available on Zenodo.

https://github.com/DidDrog11/map-liberator

## Availability and Future Directions

Map Liberator is open-source and released under the MIT License, permitting broad reuse and modification. The source code is available on GitHub (Simons [2025] 2026) with a hosted version available at shinyapps.io (“Map Liberator” n.d.). The specific version (Alpha) used for this manuscript and all future releases will be deposited in Zenodo.

I plan to expand the tool’s capabilities in three key areas:

1. To address the limitations of global datasets, future versions will allow users to upload custom shapefiles (.shp or .gpkg). This is critical for users working with non-standard geographies, such as Health Zones in the Democratic Republic of the Congo.
2. While currently manual, the selection process could be enhanced using Computer Vision (CV). We are exploring integration with opencv to suggest potential polygons based on colour matching between the reference image and the map canvas, shifting the user’s role from digitiser to validator.
3. To further support FAIR principles, I aim to implement direct export to GeoJSON and standard exchange formats compatible with DHIS2, facilitating the direct integration of extracted data into national health information systems.

Map Liberator provides a low-barrier, cost-effective solution for field epidemiologists and researchers to liberate spatial data from static reports, supporting the implementation of FAIR principles in epidemiologic research and practice (Wilkinson et al. 2016; García-Closas et al. 2023).

## Acknowledgements

Funding was provided by the joint NSF-NIH-NIFA Ecology and Evolution of Infectious Disease Award #2208034 in partnership with Research and Innovation (UKRI) Biotechnology and Biological Sciences Research Council (BBSRC) Award BB/X005364/1.

## Data availability

All data used in this manuscript are available from the Nigeria Centre for Disease Control and Prevention (Nigeria Centre for Disease Control and Prevention n.d.). The extracted data are available as supplementary material. All code to produce the tool are available on GitHub and Zenodo.

